# Phenome-wide genetic correlation analysis and genetically-informed causal inference of amyotrophic lateral sclerosis

**DOI:** 10.1101/2022.09.27.22280414

**Authors:** Salvatore D’Antona, Gita A Pathak, Dora Koller, Danilo Porro, Claudia Cava, Renato Polimanti

**Affiliations:** Institute of Bioimaging and Molecular Physiology, National Research Council, Milan, Italy; Division of Human Genetics, Department of Psychiatry, Yale University School of Medicine, New Haven, Connecticut, USA; Veterans Affairs Connecticut Healthcare System, West Haven, Connecticut, USA; Department of Genetics, Microbiology and Statistics, University of Barcelona, Barcelona, Catalonia, Spain

## Abstract

Leveraging genome-wide association statistics generated from a large study of amyotrophic lateral sclerosis (ALS; 29,612 cases and 122,656 controls) and UK Biobank (UKB; 4,024 phenotypes, up to 361,194 participants), we conducted a phenome-wide genetic correlation analysis of ALS and identified 46 genetically-correlated traits, such as fluid intelligence score (*r*_*g*_ = −0.21, p = 1.74×10^−6^), “spending time in pub or social club” (*r*_*g*_ = 0.24, p = 2.77×10^−6^), non-work related walking (*r*_*g*_ = −0.25, p = 1.95×10^−6^), college education (*r*_*g*_ = −0.15, p = 7.08×10^−5^), “ever diagnosed with panic attacks (*r*_*g*_ = 0.39, p = 4.24×10^−5^), and “self-reported other gastritis including duodenitis” (*r*_*g*_ = 0.28, p = 1.4×10^−3^). To assess the putative directionality of these genetic correlations, we conducted a latent causal variable analysis, identifying significant genetic causality proportions (gĉp) linking ALS to seven traits. While the genetic component of “self-reported other gastritis including duodenitis” showed a positive causal effect on ALS (gĉp = 0.50, p = 1.26×10^−29^), the genetic liability to ALS is potentially causal for multiple traits, also including a positive effect on “ever being diagnosed with panic attacks” (gĉp = 0.79, p = 5.011×10^−15^) and inverse effects on “other leisure/social group activities” (gĉp = 0.66, p = 1×10^−4^) and prospective memory result (gĉp = 0.35, p = 0.005). Our subsequent Mendelian randomization analysis indicated that some of these associations may be due to bi-directional effects. In conclusion, this is the first phenome-wide investigation of ALS polygenic architecture, highlighting the complex pleiotropy linking this disorder with different health domains.

## Introduction

Amyotrophic lateral sclerosis (ALS) is a complex neurological disorder due to the degeneration of the upper and lower motor neurons at the bulbar and spinal level (1), with an age of onset ranging between 50 and 65 years of age (2). Worldwide, ALS incidence is 4.5 cases per 100,000 person-year (3). The disease is characterized by initial muscle weakness that worsens to atrophy, preventing the patient from breathing and swallowing, and eventually causing death (4). Indeed, half of ALS patients die in the first three years after diagnosis of the disease and only 20% of them survive in the following seven years (5). Familial forms of ALS are due to rare mutations in genes such as *TAR DNA Binding Protein* (*TARDBP*), *Superoxide Dismutase 1* (*SOD1*), *FUS RNA Binding Protein* (*FUS*), and *C9orf72-SMCR8 Complex Subunit* (*C9orf72*) (6–9). However, more than 90% of ALS cases are not due to monogenic forms, but they are likely due to a complex genetic architecture characterized by the additive effect of many risk loci with small individual effects. To date, the largest ALS genome-wide association study (GWAS) investigated 29,612 cases and 122,656 controls, identifying 15 risk loci (10). These ALS-associated variants were shared with other neurologic traits and diseases but their effect on the transcriptomic regulation of brain regions and cell types appears to be distinct from other brain disorders (10). Beyond gene discovery, genome-wide association statistics can be used to examine the pleiotropy of ALS with other human traits and diseases. Indeed, study designs based on phenome-wide screening are invaluable tools to generate novel hypotheses regarding the genetically shared risk factors and comorbid conditions of ALS (11). In particular, genetic correlation (*r*_*g*_) analysis can provide insights into the biological pathways and/or the causality relationships linking ALS to different health domains (12). However, correlation does not prove causation. Accordingly, several methods have been developed to perform causal inference analyses using genetic information as an anchor. Indeed, while observational epidemiological studies can be affected by confounding, reverse causation, and various biases, we can use the association of genetic variants with human traits and diseases to circumvent some of these biases and generate more reliable evidence regarding the cause-effect relationships underlying complex comorbidities. Many causal inference analyses have used the two-sample Mendelian randomization (MR) approach to test the causal effect of the risk factor (i.e., exposure) on the outcome using the associations of genetic variants (i.e., instrumental variable) with exposure and outcome from two sources (13). Another established method to perform genetically informed causal inference analyses is the latent causal variable (LCV) approach which is based on the assessment of a latent variable that mediates the *r*_*g*_ between two traits of interest (14). In the present study, leveraging the genome-wide association statistics from the largest ALS study (10), we conducted a phenome-wide *r*_*g*_ analysis with respect to 957 traits available from the UK Biobank (UKB) (15). Considering traits genetically correlated to ALS, we performed causal inference analyses using LCV and MR approaches. These methods are based on different assumptions and effects consistent between them are less likely to be biased by violations of the assumptions underlying each model. We considered results consistent across these two methods to be reliable evidence of possible cause-effect relationships related to ALS pathogenesis.

## Materials and Methods

To investigate the genetic overlap and causal relationships linking ALS to other human traits and diseases, we designed the present work by applying complementary analytic approaches to multiple large-scale genome-wide datasets (Supplemental Fig. 1). This study was conducted using genome-wide association statistics generated by previous studies. Owing to the use of previously collected, deidentified, aggregated data, this study is exempt from institutional review board approval.

### Data Sources

We investigated data from the largest GWAS of ALS (10) that included 29,612 cases and 122,656 controls (91% of European descent and 9% of East Asian descent) (10). With respect to other traits and diseases, we used genome-wide association statistics generated from the UKB (15). This cohort includes approximately 500,000 individuals (54% females) from the United Kingdom with an age at recruitment between 37 and 73 (15). UKB participants were assessed for a wide range of outcomes including diet, cognitive function, work history, health status, and other relevant phenotypes. Genome-wide data were also generated in this cohort. Details regarding UKB phenotypic and genetic assessment have been previously described (15). In our analysis, we used genome-wide association statistics generated from the analysis of 361,194 UKB participants of European descent. Specifically, we analyzed 4,024 phenotypes, including traits and diseases related to different health domains (Supplemental Table 1). Details regarding the quality control and the association analysis are available at https://github.com/Nealelab/UK_Biobank_GWAS. Briefly, the genome-wide association analysis was conducted using regression models available in Hail (available at https://github.com/hail-is/hail) and including the top-20 within-ancestry principal components, sex, age, age^2^, sex×age, and sex×age^2^ as covariates. ALS and UKB genome-wide association statistics were processed by removing variants with a minor allele frequency (MAF) < 1%. With respect to UKB genome-wide association statistics generated from case-control phenotypes, we also removed variants with a minor allele count < 20 in the smaller group between cases and controls.

### SNP-based Heritability and Genetic correlation

We calculated the SNP-based heritability for ALS using genome-wide data available from a previous study (10) and for the UKB phenotypes (Supplemental Table 1). For this analysis, we used the linkage disequilibrium score regression (LDSC) (16) applying the pre-computed LD scores based on 1000 Genomes Project reference data on individuals of European ancestry (available at https://github.com/bulik/ldsc). We decided to use European-ancestry LD scores, because the ALS GWAS was generated from a sample including > 90% individuals of European descent. In line with the recommendations of the LDSC developers (16), we tested the *r*_*g*_ of ALS with UKB traits with SNP-based heritability z-score > 4. To account for multiple testing, we considered statistically significant *r*_*g*_s as those surviving a 5% false discovery rate (FDR) correction (FDR Q value < 0.05).

### Genetically-Informed Causal Inference Analysis

To identify causative effects underlying the ALS *r*_*g*_s observed, we applied the LCV approach (14). Under the assumption of a single effect-size distribution, LCV examines the presence of a single latent trait between genetically correlated traits to estimate a genetic causality proportion (gĉp), ranging from 0 to 1. Values near zero indicate partial causality, while values approaching 1 indicate full causality (14). Negative and positive gĉp values reflect the direction of the putative causal effect (i.e., phenotype #1 → phenotype #2 and phenotype #2 → phenotype #1, respectively). To avoid confusion, we describe the LCV effects oriented in accordance with positive gĉp values. Information regarding whether the LCV effect is positive or negative is provided by the LCV genetic correlation estimate (rho). The LCV analysis was performed in R using the European-ancestry LD scores.

To validate the LCV results with an independent method, we conducted a bidirectional two-sample MR analysis. Indeed, LCV and MR approaches are based on different modeling approaches. Differently from LCV method where causal effects are estimated using genome-wide information, MR approach infers putative causal relationships between exposure and outcome using a limited number of SNPs as instrumental variables (17). Specifically, MR assumptions are (i) the instrumental variables (genetic variants) are associated with the exposure, (ii) the genetic variants are not associated with confounders related to exposure and outcome, and (iii) the genetic variants affect the outcome only through the exposure factor (18).

In the present study, the MR analysis was conducted using the TwoSampleMR package (19) within the R environment. For each MR test, we defined the instrumental variable based on LD-independent variants (r^2^ < 0.001 within a 10,000-kilobase window) based on the 1000 Genomes Project Phase 3 reference panel for European populations. The clumping procedure was conducted considering a p value of 10^−5^ for the exposure GWAS. Similar to previous MR studies (20–23), we used this threshold to maximize the statistical power of the analyses performed. The analysis was conducted using five MR methods: inverse variance weighted (IVW), MR-Egger, simple mode, weighted median and weighted mode (24). These permitted to assess the MR effects, considering the sensitivities unique to each of the stated methods. We referred to the IVW method as the primary method due to its higher statistical power (25). As recommended for genetically-informed causal inference analyses (26), we avoided inference based simply on p value thresholds. The direction and strength of the effects estimated via LCV and MR analyses, together with the corresponding p values, were considered to better reflect the spectrum of evidence related to the LCV and MR results. To assess possible biases due to potential confounders (e.g., horizontal pleiotropy and heterogeneity), we performed multiple sensitivity analyses, including MR-PRESSO (Pleiotropy RESidual Sum and Outlier) global test (27), heterogeneity test (28), and the MR-Egger intercept analysis (29).

## Results

Applying the LDSC method, we calculated SNP-based heritability for ALS based on a previously-generated GWAS meta-analysis (10) and phenotypic traits available from UKB (see Data Sources). For ALS, the SNP-heritability on the observed scale was 3% (observed-scale SNP-h^2^ = 0.0357, SE = 0.0039, z = 9.15). For the 4,024 traits available from UKB, we estimated the SNP-based heritability and considered 957 phenotypes with a SNP-based heritability z-score > 4 (Supplemental Table 2) for further analyses as recommended by the LDSC developers (9). These included a wide range of outcomes related to different phenotypic domains such as “qualification: college or university degree” (SNP-h^2^ = 0.16, SE = 0.0045, z = 35.56), body mass index (SNP-h^2^ = 0.23, SE = 0.0068, z = 33.82), forced expiratory volume in one second (FEV1; SNP-h^2^ = 0.177, SE = 0.006, z=29.5), and hand grip strength (left hand; SNP-h^2^ = 0.11, SE = 0.0039, z = 28.21). The genetic correlation analysis of these traits with ALS identified evidence of genetic overlap surviving multiple testing correction for 46 of them (FDR Q value < 0.05; Figure 1; Supplemental Table 3). Ten of the ALS significant genetic correlations were related to traits derived from cognitive function tests. Specifically, ALS was genetically correlated with fluid intelligence score (*r*_*g*_ = −0.21, p = 1.74×10^−6^) and items related to its assessment (e.g., number of fluid intelligence questions attempted within time limit, *r*_*g*_ = −0.29, p = 3.09×10^−6^). Additionally, ALS was positively genetically correlated with two items related to the prospective memory test (i.e., duration screen displayed, *r*_*g*_ = 0.28, p = 1.21×10^−7^; number of attempts, *r*_*g*_ = 0.28, p = 3.63×10^−5^) and one related to the pair-matching test (time to complete round, *r*_*g*_ = 0.15, p = 1×10^−4^). Four ALS genetic correlations were related to leisure and social activities where ALS was positively genetic correlated with spending time in pub or social club (*r*_*g*_ = 0.24, p = 2.77×10^−6^) and negatively genetically correlated with being part of religious group (*r*_*g*_ = −0.22, p = 4.03×10^−5^), attending adult education classes (*r*_*g*_ = −0.24, p = 2.83×10^−5^), and other group activities (*r*_*g*_ = −0.31, p = 1.87×10^−6^). We also identified two FDR-significant genetic correlations related to non-work related transportation choices where ALS was negatively genetically correlated to walking (*r*_*g*_ = −0.25, p = 1.95×10^−6^) and using public transportation (*r*_*g*_ = −0.20, p = 6×10^−4^). With respect to education, ALS was positively genetically correlated with not having any qualification (*r*_*g*_ = 0.13, p = 1.1×10^−3^) and negatively genetically correlated with having college education (*r*_*g*_ = −0.15, p = 7.08×10^−5^), secondary education (*r*_*g*_ = −0.16, p = 5.08×10^−5^), and other professional qualifications (*r*_*g*_ = −0.20, p = 1×10^−4^). We also observed that ALS was genetically correlated with multiple mental health outcomes such as “ever thought that life was not worth living” (*r*_*g*_ = −0.21, p = 2×10^−4^), “ever contemplated self-harm” (*r*_*g*_ = −0.29, p = 2×10^−4^), and “ever diagnosed with panic attacks” (*r*_*g*_ = 0.39, p = 4.24×10^−5^). With respect to physical health, we identified genetic correlations related to self-reported other gastritis including duodenitis, *r*_*g*_ = 0.28, p = 1.4×10^−3^). Finally, we also observed that ALS was negatively genetically correlated to items related to participation in the UKB dietary questionnaire: acceptance of the invitation to complete online 24-hour recall dietary questionnaire (*r*_*g*_ = −0.32, p = 1.02×10^−7^) and number of diet questionnaires completed (*r*_*g*_ = −0.27, p = 2×10^−4^).

**Fig 1.**
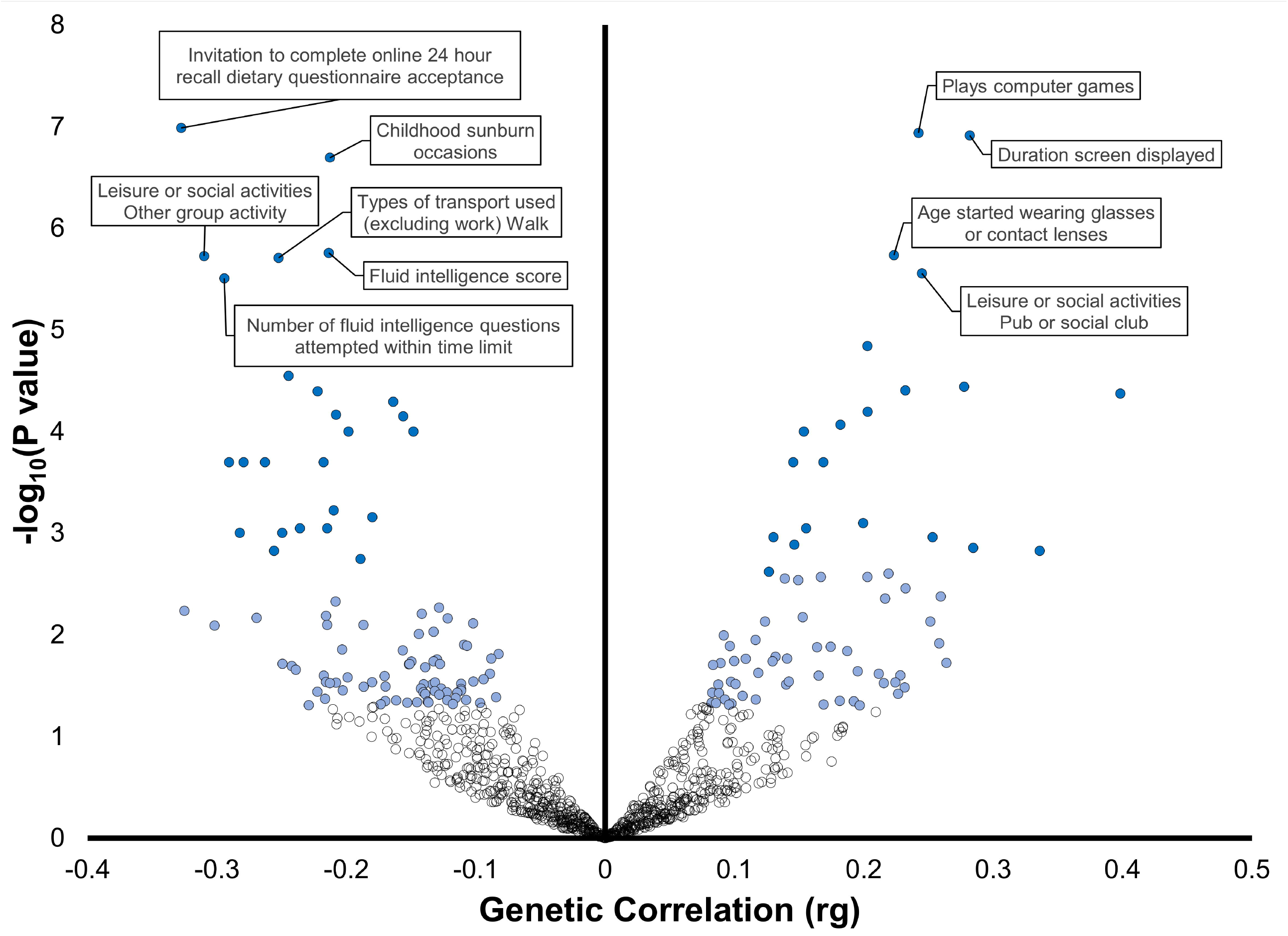
Phenome-wide genetic-correlation analysis. The color of the dots corresponds to the significance strength of the genetic correlations (rg): white (p>0.05), light blue (p<0.05), and blue (FDR q<0.05). Phenotype labels are included for the top 10 results.

To test causal effects and shared genetic mechanisms underlying traits genetically correlated with ALS, we performed a genetically-informed causal inference analysis using the LCV approach (14). After FDR multiple testing correction accounting for the number of phenotypes tested (N = 46, p < 0.05), we identified putative causal effects linking ALS to seven of the genetically-correlated traits (Figure 2; Supplemental Table 4). Only in one instance, the genetic liability of a trait (i.e., on reporting a diagnosis of “other gastritis including duodenitis”) showed a positive causal effect on ALS (gĉp = 0.50, p = 1.26 × 10^−29^). The other LCV results were related to putative causal effects of the genetic liability to ALS on other traits. Three effects were related to two traits that were positively genetically correlated with ALS (LCV rho>0): ever being diagnosed with panic attacks (gĉp = 0.79, p = 5.011×10^−15^), duration screen displayed in the prospective memory test (gĉp = 0.49, p = 6.309×10^−7^), and number of attempts in the prospective memory test (gĉp = 0.42, p = 0.001). Three effects were related to traits negatively genetically with ALS (LCV rho<0): number of diet questionnaires completed (gĉp = 0.67, p = 7 ×10^−4^), other leisure/social group activities (gĉp = 0.66, p = 1×10^−4^), and prospective memory result (gĉp = 0.35, p = 0.005).

**Fig 2.**
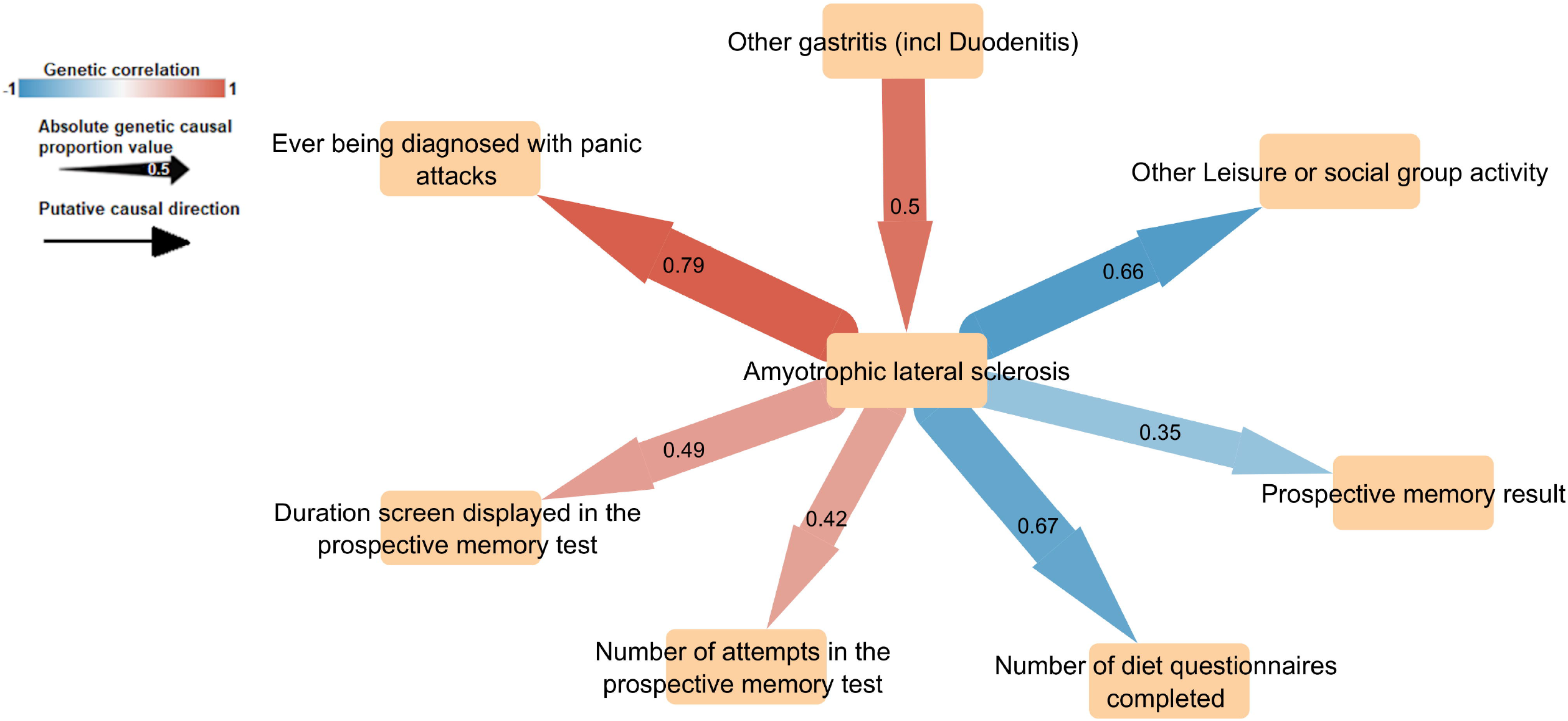
Putative causal effects linking amyotrophic lateral sclerosis with genetically-correlated traits phenotypes using the latent causal variable (LCV) approach. The color of arrows refers to the genetic correlation, while the width reflects the genetic causal proportion (gĉp), as an absolute value. The gĉp absolute values are indicated on each arrow.

To further validate the causal effects identified using the LCV approach, we bidirectional two-sample MR analysis (Supplemental Table 5). LCV and MR are based on different assumptions (14, 18). Accordingly, comparing the results obtained from these two methods permitted us to gain additional insights into the pleiotropic mechanisms linking ALS to its genetically-correlated traits. Among the causal effects identified by the LCV, we identified a consistent MR result with respect to a very small effect of the genetic liability to other leisure/social group activities on ALS (IVW beta = −0.006, SE = 0.002). Conversely, our MR analysis showed a bidirectional relationship between the duration of screen displayed in the prospective memory test and ALS (duration screen displayed → ALS: IVW beta = 0.230, SE = 0.09; ALS → duration screen displayed: IVW beta = 0.033, SE = 0.01), between the number of attempts in the prospective memory test and ALS (number of attempts → ALS: IVW beta = 0.59, SE = 0.26; ALS → number of attempts: IVW beta = 0.016, SE = 0.003) and between the prospective memory result and ALS (prospective memory result → ALS: IVW beta = 0.51, SE = 0.22; ALS → prospective memory result: IVW beta = 0.017, SE = 0.005). We also observed a significant effect of the genetic liability to the number of diet questionnaires completed on ALS (IVW beta = −0.32, SE = 0.14). This effect was characterized by heterogeneity among the effects of the variants included in the instrumental variable (IVW Q = 39.4, df = 25, p = 0.033). However, we did not identify an effect in the opposite direction (IVW beta = −0.013, SE = 0.009) or bias due to horizontal pleiotropy (MR-Egger intercept = −0.0122, p = 0.248).

## Discussion

ALS is a complex neurological disorder related to the degeneration of the upper and lower motor neurons (1, 2). Its sporadic form, the most common, is due to a complex genetic architecture characterized by the additive effect of many risk loci with small individual effects. In the present study, we used genome-wide association statistics to investigate systematically the pleiotropy linking ALS with other human traits and diseases. Through a phenome-wide screening approach, we generated new insights into putative causal effects and genetic mechanisms shared between ALS and other health outcomes. Our genetic correlation analysis showed that the polygenic architecture of ALS is correlated with traits related to different domains. In line with the enrichments for brain regions and cells observed in the ALS GWAS (10), we observed genetic correlations of ALS with multiple traits related to cognitive function and educational attainment. Previous GWAS of educational attainment and related phenotypes showed how their genetic effects have a pattern of enrichments related to brain regions and cells with stronger evidence related to neurons rather than to astrocytes and oligodendrocytes (30). With respect to cognitive function, we observed multiple phenotypes related to fluid intelligence (i.e., the ability to generate, transform, and manipulate different types of novel information in real time) and prospective memory (i.e., the ability to remember to carry out intended actions in the future). Additionally, our genetically-informed causal inference analysis showed that the genetic liability to ALS may affect these two aspects of cognitive function. ALS patients can manifest a series of cognitive impairments, including abstract reasoning (31). There is still a debate whether cognitive impairment is an early ALS symptom, or a condition related to the long-term consequence of the disease progression (31, 32). Our findings indicate that ALS polygenic risk could have a direct effect on cognitive function, supporting that cognitive impairment is an early symptom of the disease.

Another domain with multiple traits genetically-correlated to ALS is related to the assessment of dietary habits. Although diet is a factor that has been reported to alter ALS risk (33), our findings appear to indicate a more complex scenario. Indeed, while ALS is genetically correlated with cheese intake, semi-skimmed mild preference, dried fruit intake, and red wine intake, we also observed a putative causal effect of the genetic liability to ALS to the “number of diet questionnaire complete”. Recent UKB studies conducted demonstrated that participation in optional components such as the diet questionnaire can be partially predicted by loci associated with educational attainment and Alzheimer’s disease (34). Additionally, dietary patterns assessed in UKB participants appear to be causally influenced by factors correlated with education (35). Considering these previous findings and our current results, we believe that the study of diet as a possible factor to modify ALS risk should carefully account for the relationship of ALS with cognitive function and educational attainment.

Several traits genetically-correlated with ALS were related to leisure or social activities. In most of the cases, there was an inverse relation where ALS genetic effects were correlated with reduced leisure or social activities in different contexts (i.e., group activities, adult education classes, and religious groups). Additionally, the genetic liability to ALS appears to have a possible causal effect on participating in leisure group activities. This observation is in line with previous evidence reporting that cognitively intact ALS patients manifest impairments in social skills, manifesting as an inability to recognize others’ emotions and a change in behavior toward apathy (36). Although our findings highlight a general pattern of reduced social activities with respect to increased genetic risk of ALS, we also observed a positive genetic correlation between ALS and leisure or social activities in pubs or social clubs, which is consistent with the genetic correlation mentioned previously with red wine intake. The relationship between ALS risk and alcohol consumption is unclear with a recent large case-control study reporting a non-significant association (37). However, similar to other conditions, the study of alcohol consumption can be biased by misreports and longitudinal changes (38). Accordingly, further studies accounting for these possible confounders are needed to understand the potential effect of alcohol consumption on ALS risk.

Another aspect highlighted by our phenome-wide analysis is the genetic overlap between ALS and mental health. Specifically, we observed genetic correlation with severe psychiatric traits including “ever thought that life was not worth living”, “ever contemplated self-harm”, and “ever diagnosed with panic attacks”. The latter genetic relationship appears to be due to a potential causal effect of the genetic liability to ALS on the risk of panic attacks. Several studies highlighted the increased risk of ALS patients to be at increased risk of being diagnosed with anxiety, depression, and other internalizing disorders (39). Our findings support that the psychiatric comorbidity of ALS could be due to shared genetic mechanisms in addition to the negative impact of ALS on the quality of life for the affected individuals.

Among the putative causal effects identified in our genetically-informed causal inference, we observed that only the genetic liability to gastritis could be associated with increased ALS risk. Although autonomic dysfunction is reported in ALS, there is limited information about gastrointestinal symptoms among the patients affected (40). However, a growing literature is pointing to the interplay among gut microbiota and the enteric neuro-immune system as a pathogenic path to ALS and other neurodegenerative diseases (41). In this context, our findings could point toward the effect of gastritis on microbiome composition (42) as a novel area for the exploration of ALS pathogenicity.

While our study generated novel insights into ALS pathogenesis, there are two main limitations to consider. Similar to the majority of large-scale genetic studies, we investigated genome-wide data generated mostly from individuals of European descent, because of the lack of large cohorts representative of other ancestry groups. Accordingly, further studies in more diverse datasets will be needed to assess the generalizability of the associations identified in the present study. Another limitation of our study is related to the MR analysis. Indeed, because of the limited number of variants reaching genome-wide significance, we had to include suggestive loci in the instrumental variables tested. This has likely limited the statistical power of our MR analysis and may have contributed to the limited concordance between LCV and MR results.

In conclusion, our phenome-wide study generated novel insights regarding the mechanisms at the basis of the pleiotropy linking ALS to different health domains. The present results i) expanded our understanding of the possible directionality linking ALS to cognitive function, ii) highlighted the need for further studies to assess the impact of alcohol consumption and dietary habits on ALS risk, and iii) opened a new possible direction in the study of ALS pathogenesis (e.g., gastritis as an ALS risk factor).

## Data Availability

All data produced in the present work are contained in the manuscript.

## Author Contributions

SDA, CC, and RP conceptualized the study design. SDA performed analysis. SDA and RP wrote the first draft. CC and RP supervised the study. GAP, DK, DP supported the analyses and contributed to revising the manuscript. All authors gave final approval on the manuscript.

## Acknowledgments

The authors would like to thank for the financial support: “INnovazione, nuovi modelli TEcnologici e Reti per curare la SLA (INTERSLA) (ID 1157625) - Call HUB ricerca e Innovazione, promoted by Lombardy Region. The project was co-funded from the 2014-2020 POR FESR resources. IBFM-CNR thanks the support received by the Italian CNR through the Short-Term Mobility program 2021. In addition, CNR investigators acknowledge the National Recovery and Resilience Plan-National Biodiversity Future Center (NBFC)-ID: CN0000033. Yale investigators acknowledge grants from the National Institutes of Health (RF1 MH132337, R33 DA047527, R21 DC018098, and T32 MH014276), One Mind, and the European Commission (H2020 Marie Sklodowska-Curie Individual Fellowship 101028810).

## Supporting information

**S1 Figure**. Workflow of the analytic steps and methods included in the present study.

**S1 Table. UKB traits investigated in the LDSC heritability analysis**.

**S2 Table. SNP-based heritability estimates of traits available from UKB with h2 z**h.2: heritability; Std Err: standard error.

**S3 Table. ALS genetic correlations surviving multiple testing correction**. rg: genetic correlation; Q value: false discovery rate q value.

**S4 Table. ASL genetic causal proportion (GCP) surviving FDR multiple testing correction (FDR q<0.05)**. The putative causal effect are oriented in accordance with positive gcp values. gcp: genetic causal proportion; rho: genetic correlation; SE: standard error.

