## Supplementary figures and images for "Phenome-wide genetic correlation analysis and genetically-informed causal inference of amyotrophic lateral sclerosis"

### Supplemental Figure 1

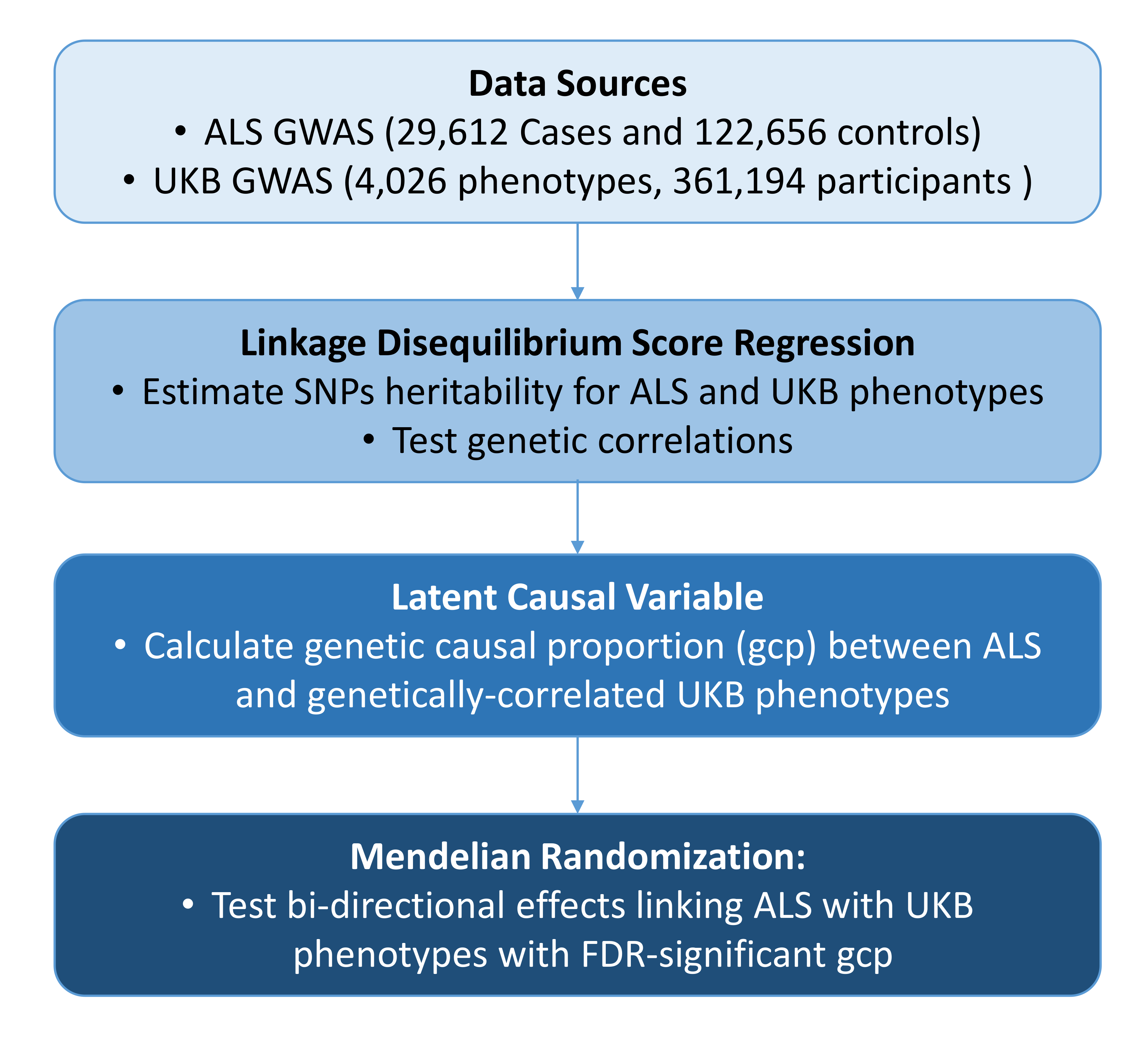
